# Bridging gaps throughout a patient’s journey with melanoma: A systematic review

**DOI:** 10.1101/2025.05.13.25327522

**Authors:** Adil Amarsi, Josh Chan, Yuan Chun Jiang, Ahmad Zobair Omar, Yasmin Meghdadi, Aashita Doshi, Alison Xie, Alyssa Wu, Joy Xu

## Abstract

**Background:** Melanoma is one of the most fatal skin cancers, with rising incidence and mortality worldwide. From diagnosis to treatment, patient experiences often involve anxiety, symptom burden, and limited access to information which profoundly impacts the quality of overall patient outcomes.

**Objective:** This systematic review aims to identify and analyze major barriers melanoma patients face throughout their healthcare journey.

**Methods:** Per PRISMA guidelines, studies were identified from PubMed, Scopus, Web of Science, Embase, and Cochrane Library, supplemented by manual hand-searching. Eligible studies focused on the experiences of melanoma patients within Western healthcare systems, addressed knowledge gaps and barriers to care throughout the patient journey, and were published in English between 2013 and 2023. Screening and extraction were conducted independently and in duplicate. Findings were synthesized based on identified themes. The methodological quality of the included studies was assessed using the GRADE criteria.

**Results:** Out of 2,257 screened articles, 183 met the inclusion criteria. Studies were categorized into four major themes: intersectionality, treatment, diagnosis/prognosis, and patient/societal burden. Commonly explored subcategories included self-examination, risk factors, and drug efficacy. This focus was driven by the emergence of novel self-examination interventions and knowledge gaps regarding risk factors and prognosis, particularly in relation to newly introduced immune checkpoint inhibitors and targeted therapies.

**Conclusions:** Melanoma patients experience significant gaps throughout their healthcare journey. Identifying areas of improvement in current practices is the first step toward developing targeted solutions that improve the patient experience and quality of life.

## INTRODUCTION

Despite significant advances in the treatment of malignant melanoma, it remains the deadliest form of skin cancer ^1–3^. With an estimated 325,000 cases and 57,000 deaths worldwide ^4^, addressing melanoma is a critical public health priority. Projections based on 2020 trends suggest that incidence and mortality could rise by approximately 50% and 68%, respectively, within the next two decades ^4^. Simultaneously, there remains a notable gap in the literature regarding the emotional well-being and distress associated with melanoma ^5,6^.

A decade after the introduction of immune checkpoint inhibitors (ICIs) and targeted therapy (TT), alongside recent enhancements in telemedicine and artificial intelligence, clinical standards for melanoma are evolving rapidly. As a result, current understandings of the melanoma patient journey necessitate a comprehensive review of recent clinical advancements, gaps in the literature, and policy shortcomings from a patient-centered perspective. In response to these considerations, a review was conducted to recognize crucial factors impacting the quality of life for melanoma patients. The objective of this study was to assess the current landscape of research on these issues and identify areas where further investigation is needed.

## METHODS

### Study Design, Inclusion Criteria, and Data Sources

A comprehensive review was conducted per Preferred Reporting Items for Systematic reviews and Meta-Analyses (PRISMA) guidelines (see Figure 1 for the PRISMA flowchart). Studies were identified by searching PubMed, Scopus, Web of Science, Embase, and Cochrane Library on December 27, 2023. The enacted search strategy in PubMed was the following, with equivalent search strategies being applied in other databases:

<u>Search strategy (applied in full text):</u> Melanoma AND (diagnos* OR treat* OR prognos* OR prevent*) AND (healthcare* OR physician* OR patient* OR manag* OR system* OR communit* OR grassroot* OR patient experience* OR experien* OR lifestyl* OR guideline* OR barrier* OR challenge* OR obstacle*)
<u>Filters:</u> Free full text, Full text, in the last 10 years, Humans, English, Child: 6-12 years, Adolescent: 13-18 years, Adult: 19+ years, Young Adult: 19-24 years, Adult: 19-44 years, Middle Aged + Aged: 45+ years, Middle Aged: 45-64 years, Aged: 65+ years, 80 and over: 80+ years.

**Figure 1:**
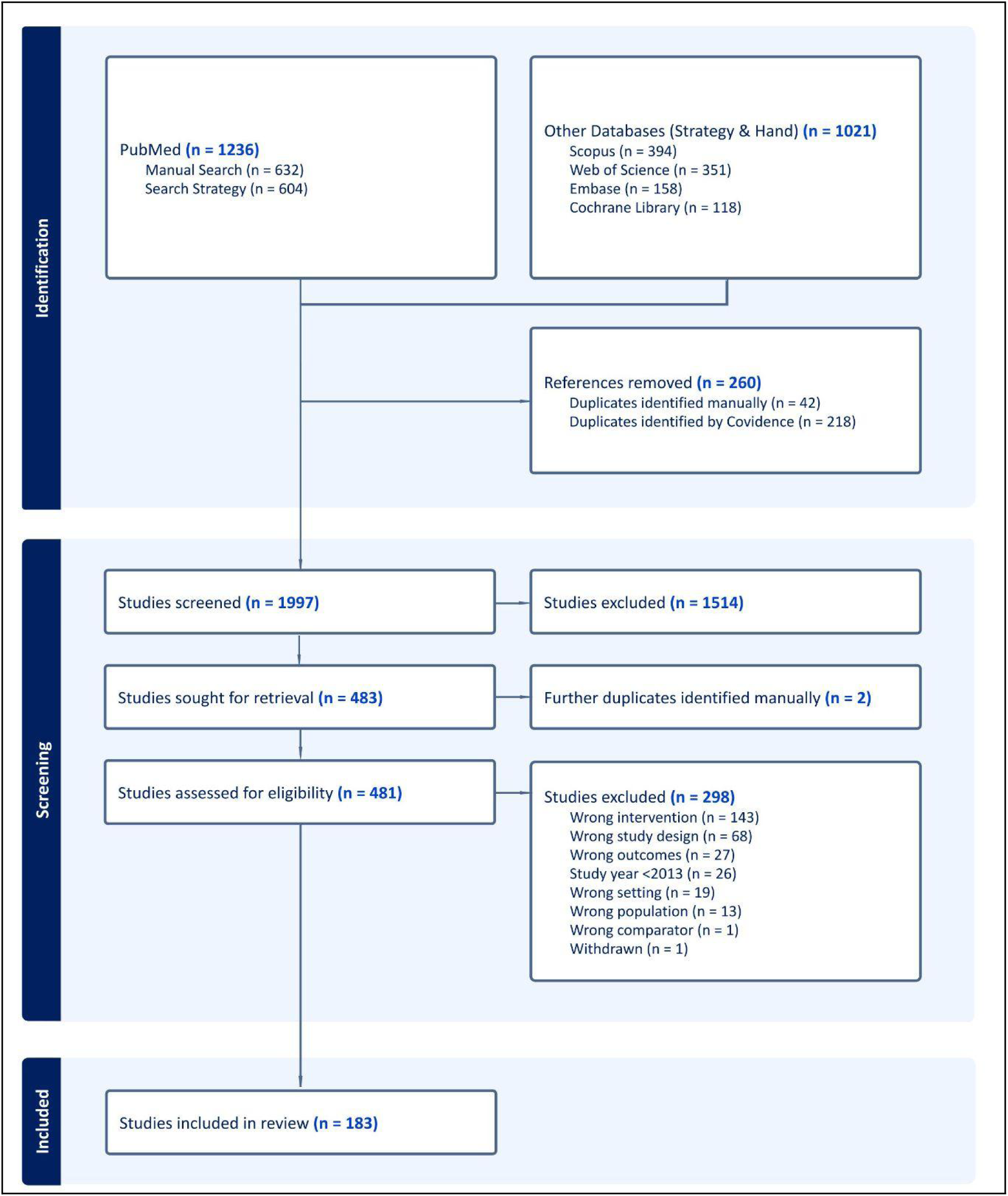
PRISMA diagram for included studies

Studies were included through duplicate independent review (see Study Selection and Data Extraction for further details) if they met the following criteria: (1) primarily focused on melanoma patients; (2) examined any diagnostic, prognostic, therapeutic or societal gaps and/or solutions for patients; (3) were published in English; and (4) were conducted between 2013 and 2023. Exclusion criteria comprised of (1) conference abstracts or posters; (2) non-peer-reviewed sources; (3) animal or *in vitro* studies; (4) studies with the majority of the data (patients or included studies) collected from South America, Africa, Asia, the Americas (excluding Canada and the US), and Europe (excluding European Union member states, the United Kingdom, and Switzerland); and (5) RCTs or observational studies solely focused on the effect of pharmaceuticals on melanoma, as this is not relevant to the research objective.

Reference lists of included studies were hand-searched for additional eligible articles. The hand-searching process was automated using Python, leveraging article titles to locate corresponding PubMed pages. For successfully retrieved articles, the similar articles section and reference lists were extracted, including the five PubMed-suggested articles and only references with valid PubMed links, thereby excluding grey literature. Articles without a PubMed page or those with ambiguous search results were excluded from the hand-search process. Additionally, articles lacking a reference list on PubMed did not have their reference list screened. The code used for automation is available upon request.

### Study Selection and Data Extraction

Five reviewers (AA, JC, YCJ, YM, and AD) performed a pilot test followed by title and abstract screening independently and in duplicate. Full-text review was also conducted independently and in duplicate. At both stages of the review, conflicts were resolved by a blinded third reviewer. Reviewers AA and JC subsequently independently piloted the data extraction form using a random sample of studies, implementing necessary revisions within Covidence based on their evaluation. Extracted data included study location, study design, population type, sample size, demographic characteristics (age, sex, and race), melanoma subtype, intervention details, outcomes, and gaps identified through the patient journey with melanoma.

### Risk of Bias Assessment

The risk of bias in the included studies was independently assessed in duplicate by seven reviewers (AA, JC, YCJ, AZO, YM, AD, and AX). To evaluate the quality and certainty of evidence, the Grading of Recommendations, Assessment, Development, and Evaluation (GRADE) tool was used. This approach examines key factors within the evidence-to-decision framework, including sequence generation, allocation concealment, blinding of participants, personnel, and outcome assessors, incomplete outcome data, selective outcome reporting, and other sources of bias.

## RESULTS

### Study Selection and Characteristics

The database search and hand-searching process yielded a total of 2,257 studies, of which 260 duplicates were removed. A total of 1,514 studies were excluded during title and abstract screening, and another 298 were excluded during full-text review. Detailed information regarding the full-text review process is provided in Figure 1. Overall, 183 studies met the inclusion criteria for data extraction and risk of bias assessment.

The 183 studies in this review included 100 cohort studies and/or retrospective analyses ^7–106^, 32 literature reviews or qualitative patient research/interviews ^107–138^, 19 systematic reviews and/or meta-analyses ^139–157^, 9 cross-sectional studies ^158–166^, 8 randomized controlled trials (RCTs) ^167–174^, and 15 mixed/other methods studies ^175–189^. Geographically, 73 studies were conducted in the United States, 27 in individual European Union countries, 26 in Canada, 12 in Australia, 6 in China, 2 in Switzerland, 2 in the United Kingdom, and 35 across multiple countries. Included Chinese studies used data collected in the US or across various included countries.

### Outcomes

The analysis of the included studies resulted in the identification of four main themes related to gaps in melanoma care: intersectionality, treatment, diagnosis/prognosis, and patient/societal burden. These themes were further divided into subcategories (Table 1). Each subcategory was analyzed, with greater emphasis placed on studies with a lower risk of bias (see Risk of Bias Assessment); however, every included study was analyzed.

**Table 1:**
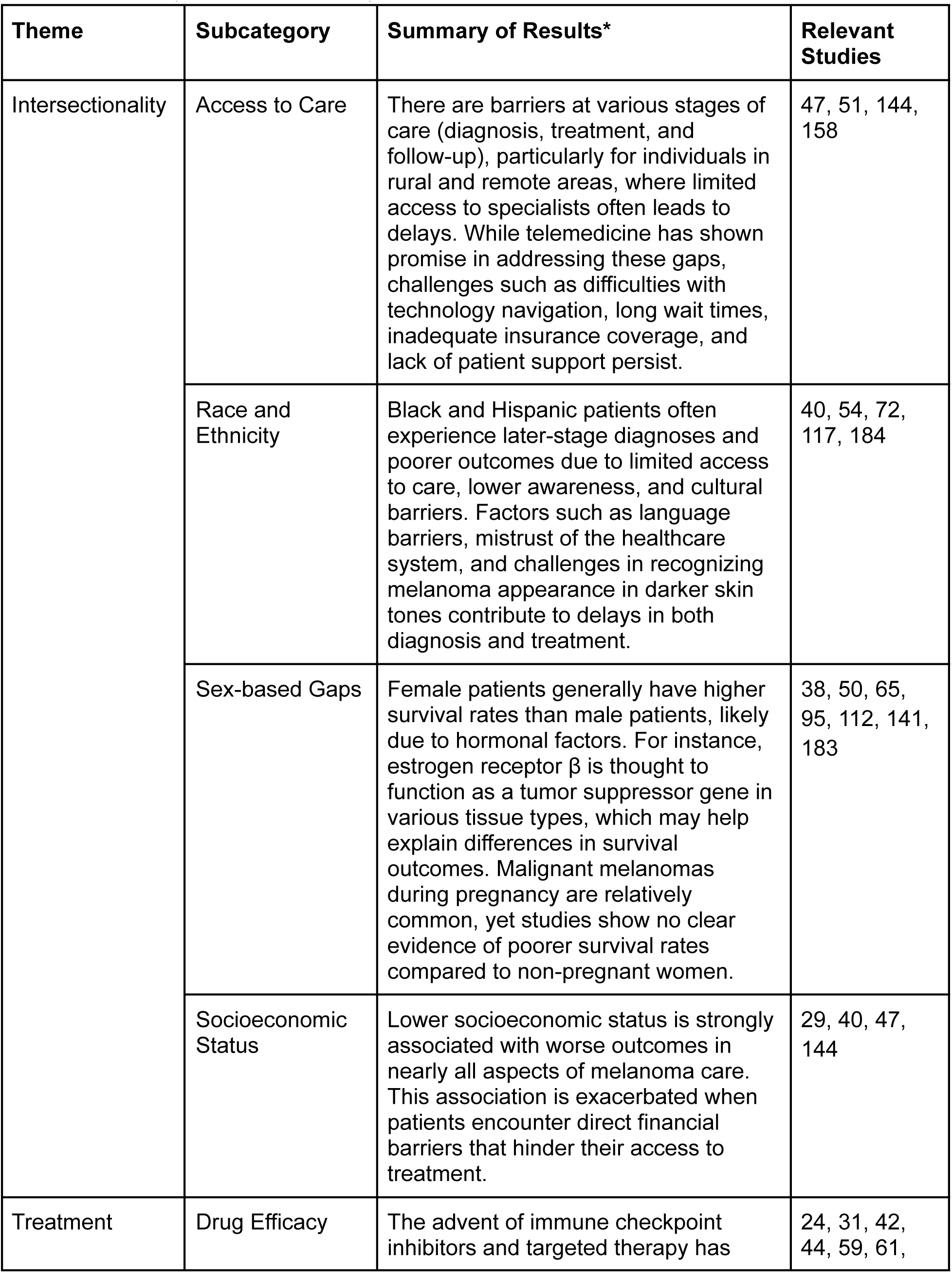
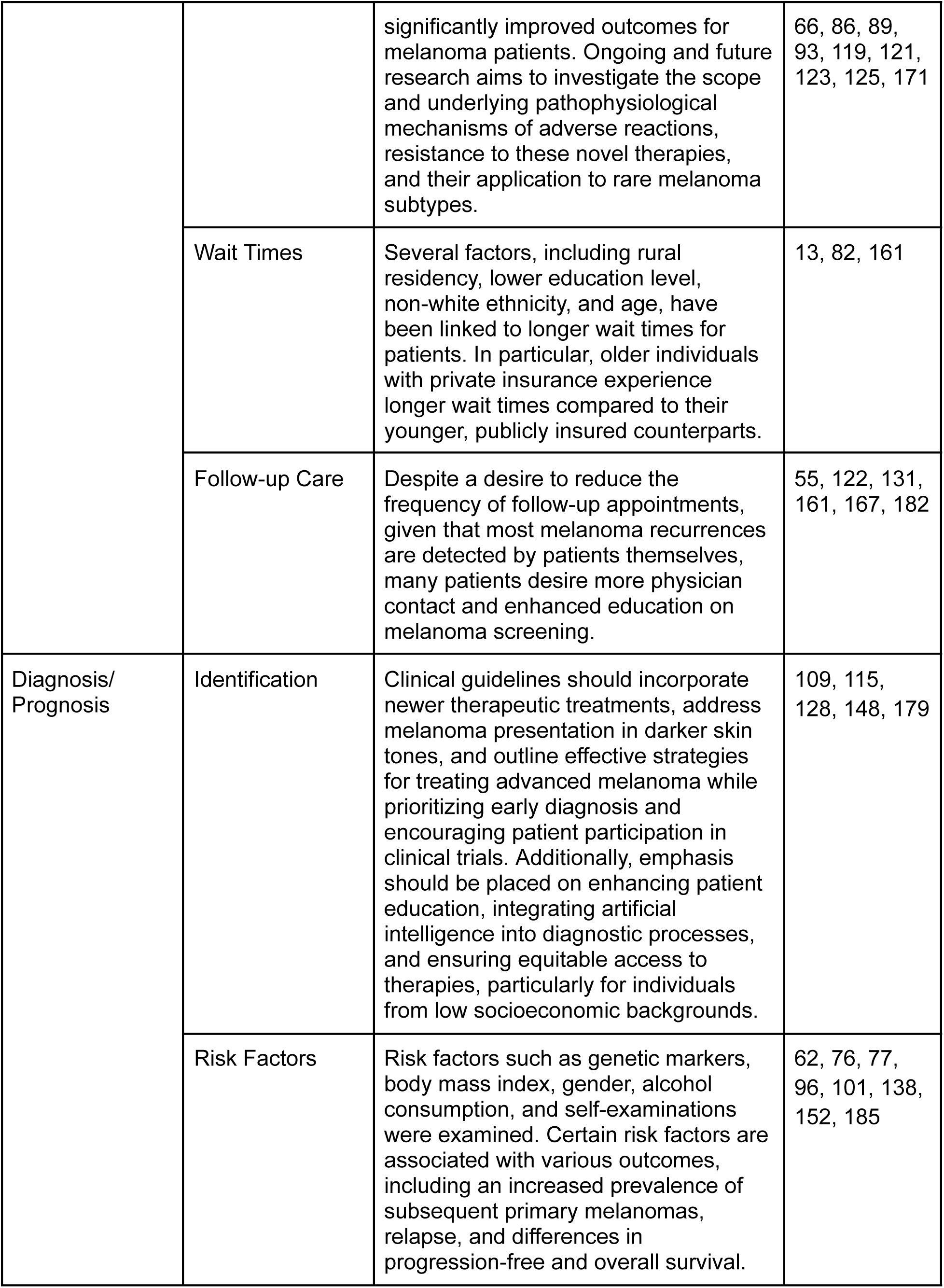
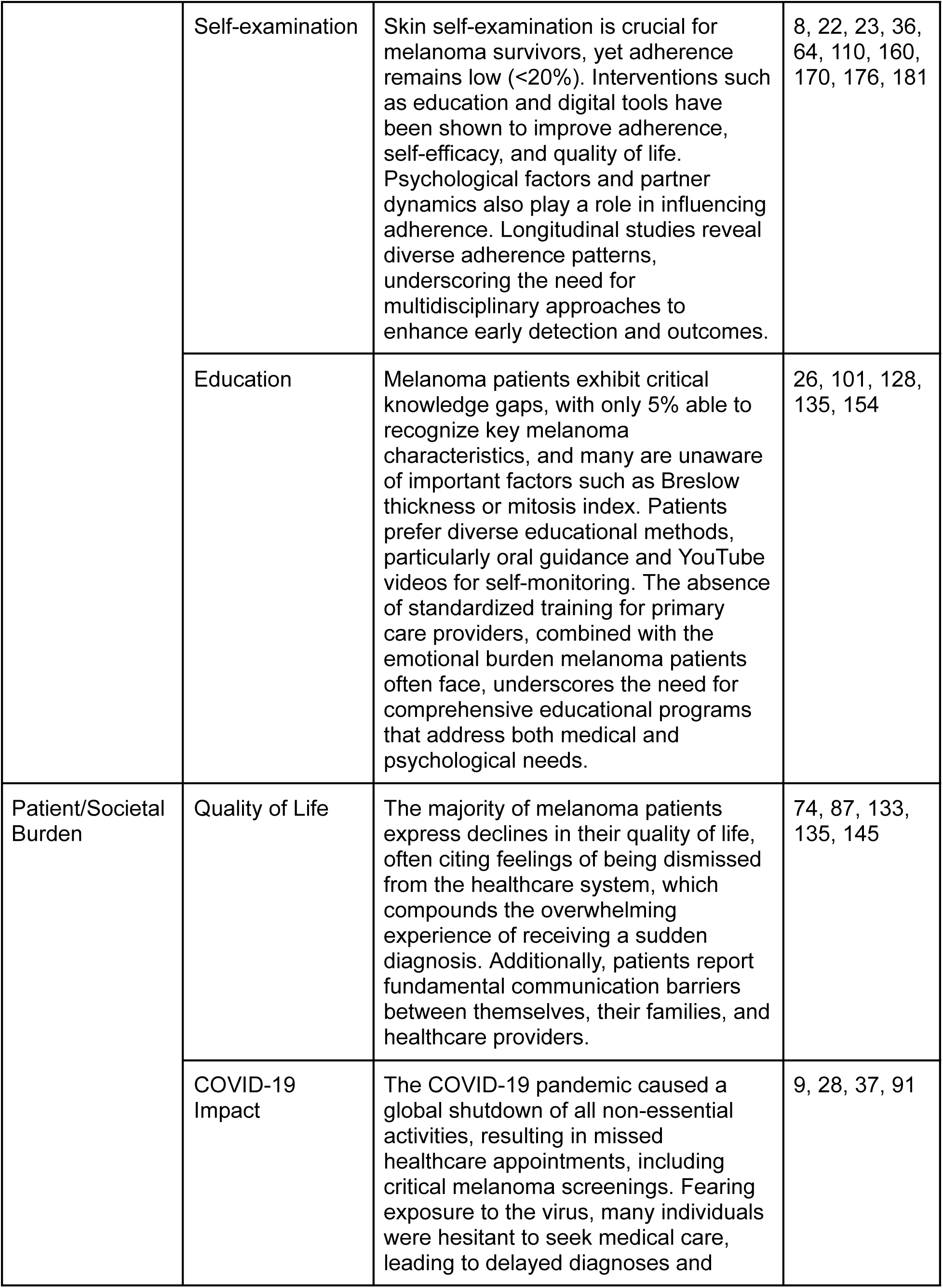
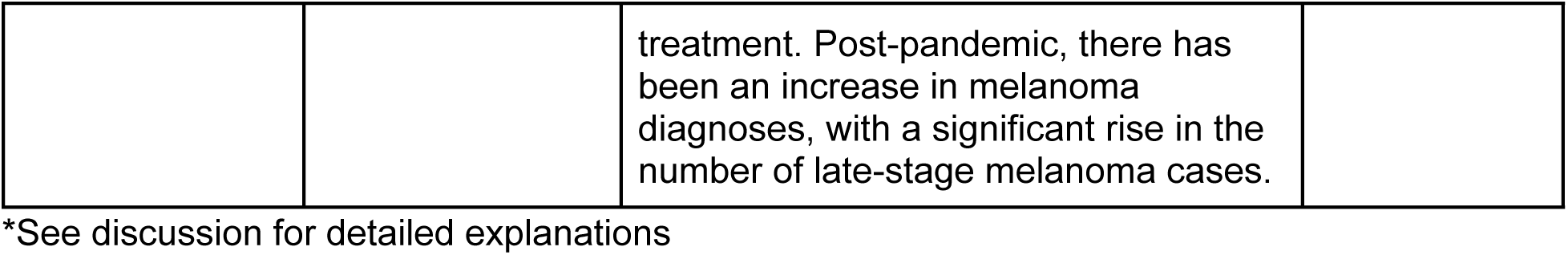
Summary of Results and Key Articles.

### Risk of Bias Assessment

The risk of bias was low in 143 studies (78.1%), moderate in 22 studies (12%), and high in 18 studies (9.8%) (Figure 2).

**Figure 2:**
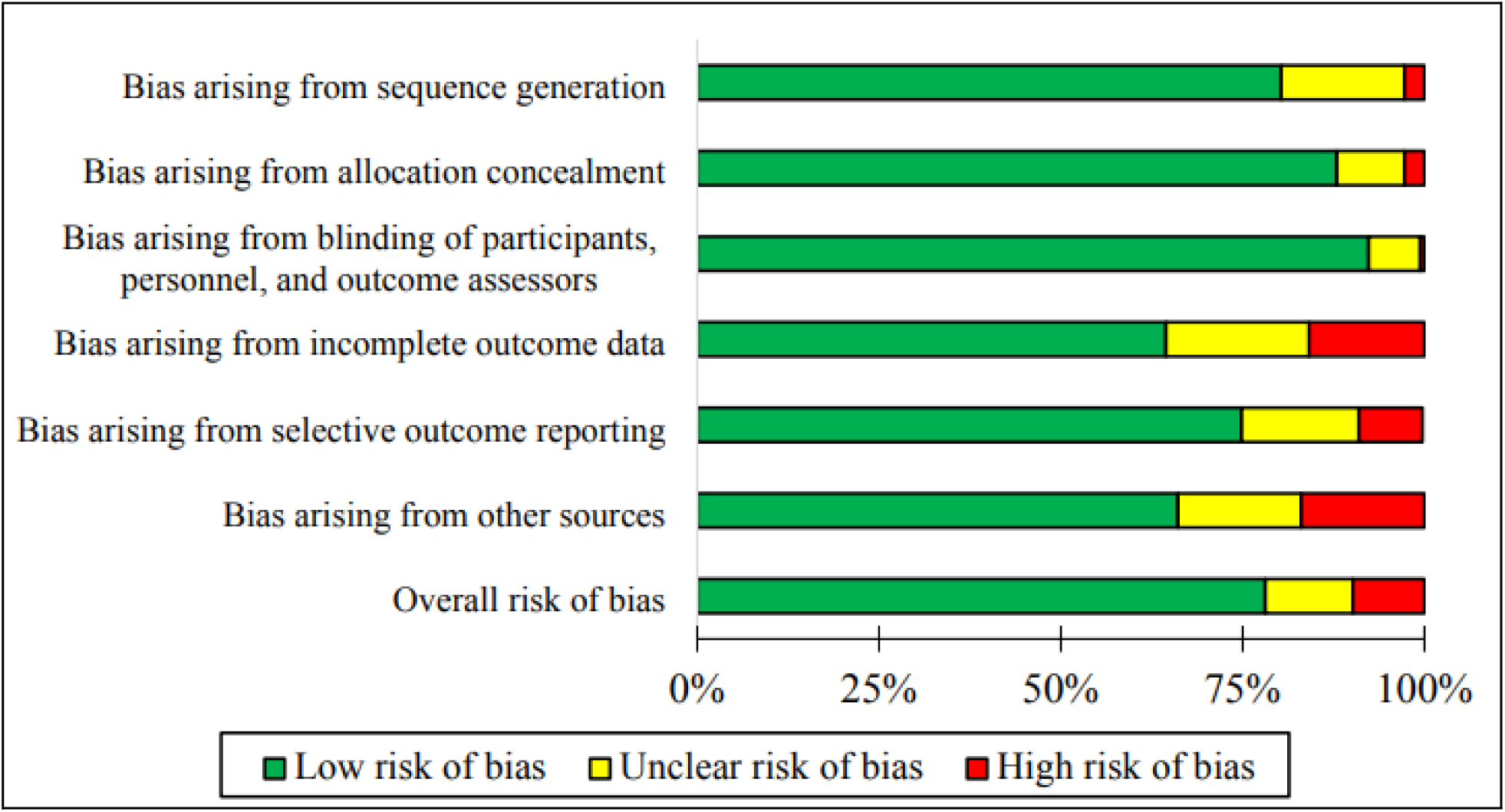
Risk of bias assessment results

## DISCUSSION

### Intersectionality

#### Access to Care

Studies examining access to care for melanoma patients reveal significant barriers at various stages of the patient journey, including specialist referral, initial diagnosis, and treatment initiation. For instance, a study investigating the use of immunotherapy for metastatic melanoma found that underinsured patients were 2.44 times more likely to receive treatment at hospitals with low immunotherapy prescribing rates ^51^. Geographic disparities were a prominent theme, with patients in rural and remote areas often experiencing delays due to limited access to dermatologists and oncologists. In a study exploring factors associated with delayed melanoma diagnosis, geographic location was found to be significantly associated with a diagnosis of advanced-stage disease ^47^. Although some studies highlighted the potential of telemedicine to mitigate these access gaps, its effectiveness is not always consistent. For example, an RCT demonstrated that teledermoscopy did not improve the sensitivity of skin cancer detection. Additionally, systemic issues such as lengthy wait times for appointments, limited specialist availability, inadequate health insurance coverage, and lack of patient navigation support were identified as key challenges. Collectively, these findings underscore the need for integrated, patient-centered approaches to ensure equitable access to healthcare throughout a patient’s journey with melanoma.

#### Race and Ethnicity

Studies focusing on race and ethnicity in melanoma care reveal significant disparities in diagnosis, treatment, and patient-reported outcomes. Racial and ethnic minorities, particularly Black (p = 0.024) and Hispanic (p < 0.001) populations were more likely to experience later-stage diagnoses and poorer outcomes compared to their White counterparts ^72^. These disparities were often attributed to lower awareness of risk factors and reduced access to care. For example, Black individuals are significantly less likely to receive surgical resection treatment for localized disease, despite its well-established benefits in improving survival rates. Cultural factors, language barriers, and mistrust of the healthcare system stemming from historical injustices further exacerbated delays in seeking and receiving care. These have contributed to a lower 5-year survival rate among Black individuals compared to their White counterparts (58.8% vs 84.8%). Furthermore, some studies noted challenges in recognizing melanoma in individuals with darker skin tones, often resulting in delayed or missed diagnoses. To address this, research suggests increasing the availability of melanoma images featuring darker skin tones. Overall, these findings emphasize the need for targeted strategies to address racial and ethnic disparities in melanoma care, ensuring that all melanoma patients receive timely and accurate care.

#### Sex-based Gaps

Differences in biological sex significantly impact the survival outcomes of melanoma patients, with female patients generally experiencing better cancer-specific survival (CSS) rates than their male counterparts. For instance, women under 45 years of age showed markedly higher CSS rates compared to men of the same age group for stage II and III melanoma. Unadjusted 3- and 5-year CSS estimates were 64.2% vs. 59.7%, and 53.5% vs. 49.9%, respectively (p ≤ 0.0001) ^43^. The literature suggests that differences in hormonal and genetic composition may be the reason for this disparity. For example, estrogen receptor β (ERβ) expression is thought to be a key modulator of tumor behavior. When estrogen binds to its receptor, it triggers tumor suppression in various tissues, including the skin. Research indicates that ERβ expression decreases as disease severity and tumor aggressiveness increase, with the highest expression found in benign cases and the lowest in malignant disease ^43^. Additionally, genomic analysis of metastatic melanoma, including whole exome sequencing, reveals a higher frequency of missense mutations in male tumor samples. Studies of UV hotspot mutations suggest these genetic differences are not primarily caused by environmental factors such as UV light exposure, but may instead reflect intrinsic genetic differences ^43^.

Pregnancy-associated melanoma (PAM) is another factor contributing to the sex-based survival gap between female and male patients. Approximately one-third of melanomas in women occur during their child-bearing years, making melanoma one of the most common malignancies during pregnancy. PAM occurring postpartum is also associated with greater tumor thickness compared to non-PAM cases, with a difference of 2.01-4.00 mm vs. 0.01-1.00 mm, respectively ^50^. Additional factors, such as the impact of PAM on mental health and emotional well-being, remain unclear and warrant further investigation.

#### Socioeconomic Status

Socioeconomic status (SES) plays a critical role in the survivability of melanoma patients at all stages of the patient journey. Often measured through factors such as income, education, occupation, and social perception, SES can also be represented indirectly by aspects such as insurance coverage. Together, these factors can severely impact one’s ability to manage and access melanoma care. Lower SES is linked to delayed diagnoses, limited access to treatment, and detection at more advanced disease stages. In particular, a study by Cortez et al. in Florida found that from 1999 to 2008, every 1% increase in poverty was associated with a 2% increase in late-stage melanoma diagnoses (p < 0.001) ^144^.

Patients with lower SES also face restricted access to treatment and care. For example, in the United States, patients with Medicaid, the country’s government-sponsored health insurance program, were significantly more likely to experience delays in surgery for therapeutic excision compared to those with private insurance. This trend was also seen in the ability to access innovative therapies and dermatologic coverage for certain cases ^144^. Additionally, studies have shown that lower levels of education are associated with a higher probability of a stage IV diagnosis. Individuals with lower levels of education may be less aware of their risk for melanoma, leading to a failure to seek regular skin examinations. Research has also found that patients without a high school diploma received fewer instructions from their physicians on how to identify signs of melanoma, were less likely to be informed about their risk of skin cancer, and less likely to have discussions about skin cancer at all ^29^. As a result of these factors, patients with lower SES face poorer overall and melanoma-specific survival rates compared to their higher SES counterparts. This underscores the urgent need for equitable healthcare policies, advocacy initiatives, and preventative or screening programs to address SES-related disparities and improve melanoma survival outcomes.

### Treatment

#### Drug Efficacy

While most melanoma patients are diagnosed with resectable stage I or II disease, outcomes for more advanced melanomas remain poor, with a 5-year overall survival (OS) of less than 19% ^123^. The introduction of ICIs and TT in 2011 revolutionized treatment for metastatic melanoma. Prior to these novel treatments, traditional chemotherapies such as dacarbazine struggled to improve patient outcomes, with median OS ranging from 5 to 11 months ^121^. ICIs and TT have improved survival, with median OS now exceeding 20 months in some cases. Ongoing research is exploring their efficacy as second-line therapies or in combination with surgery and radiation ^61,121^. However, due to their relatively recent introduction, optimizing dosages, managing adverse effects, and identifying prognostic factors remain areas of further investigation.

ICIs work by enhancing the immune response against melanoma, primarily targeting specific immune receptors ^121^. In practice, combination therapies have shown superior efficacy compared to monotherapy. For instance, phase I trials reviewed by Michielin et al. demonstrated that the ipilimumab and nivolumab (N/I) combination achieved an 85% 1-year OS compared to 46% and 73% for ipilimumab and nivolumab, respectively ^121^. Beyond aiming to improve survival outcomes, current research on ICI combination therapy focuses on reducing adverse events (AEs) for patients. The Checkmate 218 program reported that 96% of ICI recipients experienced AEs, which most commonly included fatigue, diarrhea, nausea, and pruritus ^44^. Severe AEs led to treatment discontinuation in up to 36% of participants. Dai et al. found that 30.5% of second-line ipilimumab recipients required specialist care post-treatment, compared to 19.5% for those receiving alternative second-line therapies. The study also found that ipilimumab patients were more likely to experience hospitalization (OR 1.81) ^24^.

To mitigate AEs while maintaining survival benefits, research is focused on optimizing ICI dosing regimens. The Checkmate 511 trial demonstrated that 33.9% of patients receiving a regimen of 3 mg/kg nivolumab and 1 mg/kg ipilimumab (3N1I) experienced fewer grade 3-5 AEs compared to 48.3% in those receiving 1 mg/kg nivolumab and 3 mg/kg ipilimumab (1N3I), while maintaining comparable survival outcomes (median progression-free survival [PFS] of 8.94 months vs. 9.92 months) ^171^. Ma et al. explored the impact of treatment discontinuation and found that among patients with similar clinical benefit responses after one or two doses, completing all doses was not significantly associated with improved PFS (p = 0.921) or OS (p = 0.965) ^59^. These findings suggest that dose optimization could improve patient quality of life while ensuring treatment efficacy. Currently, AEs are managed through concurrent medications or treatment discontinuation when necessary, but further research is required to refine dosing strategies.

Alongside ICIs, TT has emerged as another novel therapy for metastatic melanoma, particularly for patients with specific genetic mutations. BRAF mutations, which are found in over 50% of melanomas, can be targeted with BRAF inhibitors such as vemurafenib, which has improved OS from 10 to 13.8 months ^119^. TT is often favored as a first-line treatment for rapidly progressing patients due to its high objective response rate (ORR), oral administration, and AEs associated with ICIs ^125^. Despite these advantages, 50% of patients receiving BRAF inhibitors experience disease progression due to resistance to TT ^121^. Studies suggest that resistance develops due to genetic mutations affecting the BRAF/MEK/ERK pathway, with up to 58% of patients exhibiting mutations such as NRAS/KRAS alterations or BRAF splice variants ^119^.

Although these advancements have improved outcomes for cutaneous melanoma, rare melanomas have seen mixed results. Melanoma of unknown primary (MUP) has benefited from novel therapies, with OS improving from 4 to 11 months in stage IV cohorts. However, some studies have reported worse TT outcomes, with a 47% 1-year survival rate compared to 56% in other studies ^89,93^. In contrast, uveal melanoma has shown poor response to both TT and ICI since BRAF mutations are less common in uveal melanoma. 83% of uveal melanomas involve activating mutations in the GNAQ or GNA11 genes ^66^. N/I combination therapy has shown promise, increasing ORR to 15.6%, but remains far less effective than in cutaneous melanoma, where ICI monotherapy alone achieves an ORR of 45% ^42^.

Brain metastases present another challenge in melanoma treatment, as patients with these metastases are frequently excluded from clinical trials, leaving knowledge gaps on the efficacy of ICIs and TT for this subgroup. Some studies have reported improved outcomes, with response rates increasing by up to 31% with novel TT. However, ICIs and TT have not significantly reduced the incidence of brain metastases compared to chemotherapy, likely due to the blood-brain barrier restricting drug penetration ^86^. Combination TT approaches have shown limited success, with 87.7% of patients experiencing disease progression and an OS of just 9.5 months ^31^.

#### Wait Times

Conic et al. identified wait times as a significant prognostic factor, with delays exceeding 90 days correlating with a 29-41% decrease in OS ^161^. Expanding on this, Baranowski et al. explored socioeconomic and health-related factors contributing to longer wait times. Their analysis revealed that non-White minorities and individuals with lower educational attainment, nonmetropolitan residency, and multiple comorbidities were more likely to experience treatment delays ^13^.

#### Follow-up Care

The MELanoma Follow-up (MELFO) study sparked discussions on reducing follow-up frequency. Since almost 75% of melanoma recurrences are detected by patients themselves and in between follow-up visits, researchers have hypothesized that reducing follow-up frequency may not compromise detection accuracy or patient care. Damude et al. demonstrated that reducing physician interactions resulted in an average cost reduction of 45% per patient ^167^. Despite this finding, studies on patient perspectives of follow-up care suggest a strong preference for physician-led detection, with significantly lower satisfaction scores in the reduced physician interaction group (p = 0.01). Lim et al. found many patients lack confidence in their ability to perform self-examinations, with only 38% feeling capable of detecting lesions ^55^. Similarly, Mitchell et al. reported widespread dissatisfaction with the current state of follow-up care, with 64.1% of respondents expressing a desire for further education and communication from their healthcare providers ^182^. Given these concerns, any modifications to follow-up protocols must account for the preference of patients for less independent follow-up arrangements.

### Diagnosis/Prognosis

#### Identification

Studies analyzing melanoma diagnosis and identification guidelines have highlighted key areas

for improvement, including enhancing early detection, addressing missed diagnostic opportunities, evaluating the efficacy of post-diagnosis follow-up protocols, and comparing international guidelines to identify more predictive classification methods. Clinical guidelines are important to account for emerging treatments and consider the presentation of melanoma in minority groups, including African American, Asian, and Hispanic populations.

Evaluations of guideline effectiveness have underscored that standardized, evidence-based guidelines can lead to earlier diagnosis and prevent disease progression. However, inconsistencies in follow-up protocols remain. While less than 5% of melanoma recurrences occur beyond five years post-treatment, studies suggest that the risk of developing subsequent primary melanoma persists for more than a decade after initial diagnosis. Thus, establishing clear guidelines that integrate time-limited clinical follow-ups with lifelong self-examinations could significantly improve relapse detection. Nonetheless, effective treatment strategies for advanced melanoma remain limited, contributing to variability in treatment recommendations and decreasing OS rates ^179^. The literature suggests that increasing patient participation in RCTs and a shift toward personalized cancer therapies can help address gaps in treatment. As research evolves, a universal, scientifically validated melanoma treatment framework may provide patients with optimal care and improved long-term survival outcomes ^179^.

Educational initiatives were also emphasized as critical for both the general public and medical professionals. Technological advancements, such as dermatoscopes, sequential digital dermoscopy imaging, and teledermatology, could lead to earlier detection and reductions in disease-specific mortality ^115^. For example, a teledermatology study found that over 50% of cases could be managed without the need for a dermatologist, leading to a 78% reduction in wait times for in-person appointments. Expanding training programs and melanoma education programs can help alleviate the burden on dermatologists while enhancing patient outcomes.

Artificial intelligence has emerged as a promising tool in melanoma diagnosis, with deep convolutional neural networks performing at diagnostic levels comparable to dermatologists ^109^. However, concerns remain regarding potential racial and socioeconomic biases embedded in AI algorithms. Since melanoma exhibits unique demographic, clinical, and genetic characteristics in African American, Asian, and Hispanic populations, including in gender prevalence and subtype, it is critical to ensure that AI models are trained on diverse datasets. Addressing these concerns is imperative for improving early detection and treatment ^148^.

While self-screenings, non-specialist detection, and new technologies offer new diagnostic opportunities, they also raise some concerns about overdiagnosis, increased healthcare costs, and uncertain survival benefits ^115^. Equal access to diagnostic and therapeutic advancements must be prioritized, particularly for individuals from low socioeconomic backgrounds, without regular physician contact or smartphone access, and those from rural areas ^115^.

#### Risk Factors

The literature has evaluated a range of risk factors associated with melanoma prognosis, treatment response, and relapse, including genetic markers, body mass index (BMI), gender, alcohol consumption, and self-examination practices. The risk of developing a subsequent primary melanoma varies significantly between individuals and is particularly high for those with two or more primary melanomas ^175^. Notably, 13.4% of patients with a high-risk primary melanoma experience disease recurrence within two years ^96^. Tumor location on the head or neck, sentinel lymph node biopsy positivity, and signs of rapid tumor growth are other key predictors of relapse ^96^.

The presence of brain and liver metastases has been associated with shorter PFS and OS following second-line treatment. High lactate dehydrogenase levels have also emerged as a clinically significant biomarker, correlating with shorter survival in patients receiving anti-PD1 immunotherapy after BRAFi/MEKi combination therapy ^185^. However, the same study emphasized that patients with high tumor burden, including brain metastases, may benefit from second-line therapy, with approximately 20% achieving an ORR exceeding 65% and another 20% experiencing disease stabilization. While the relationship between alcohol consumption and melanoma risk remains unclear, evidence suggests that downregulation and inactivation of ethanol-metabolizing enzymes may play a role in preventing melanoma initiation and progression ^138^. Further research incorporating molecular and translational analyses is needed to identify additional predictive and prognostic biomarkers. Advancing the study of biomarkers will enhance treatment decision-making and improve outcomes, ultimately facilitating a more personalized approach to melanoma management.

BMI has been significantly investigated as a prognostic factor, with many studies reporting an association between higher BMI and improved survival outcomes. McQuade et al. found that obese patients had a median OS of 33 months compared to 19.8 months for those with a normal BMI. This trend was observed in both ICI and TT recipients, though the survival benefit was only significant in men ^62^. Naik et al. further corroborated this finding, reporting a median PFS of 683 days in overweight and class I obese patients compared to 135-163 days in those with a BMI below 25 ^65^. However, Donnelly et al. attempted to replicate these findings and found no significant difference in most survival outcomes, except for a positive correlation between overweight (p = 0.02) and obese (p = 0.01) ICI recipients and improved PFS ^30^. Interestingly, the authors also found a correlation between weight gain during treatment and worse disease response, suggesting that actively increasing weight during treatment does not improve outcomes ^30^. Finally, Rutkowski et al. found no significant impact of BMI on disease control rate, PFS, and OS in patients with metastatic melanoma receiving immune ICIs ^77^. Given these conflicting findings, further research is needed to establish a consensus on the implications of BMI on melanoma prognosis and treatment response.

The impact of melanoma screenings on prognosis has also been explored. In one study, community screening initiatives led to a 79% increase in melanoma diagnoses ^101^. However, there was no significant increase in rates of skin surgeries or dermatology visits. These findings suggest that while community screenings can enhance melanoma detection, their broader impact on patient outcomes remains uncertain. To meaningfully improve prognosis following a melanoma diagnosis, efforts should focus not only on expanding screening initiatives but also on increasing access to affordable care and specialized physicians.

#### Self-examination

Skin self-examination (SSE) is an important early detection practice for melanoma survivors and patients at risk; however, its implementation remains hindered by various challenges. Despite professional recommendations, patient adherence remains low, with fewer than 25% of survivors conducting monthly checks and only 14.2% performing a thorough full-body examination ^23,36^. However, novel interventions are showing promise in improving SSE practices. For example, educational programs have increased self-efficacy by 23%, with benefits lasting up to a year ^176^. Additionally, digital tools such as tablet-based support systems have increased SSE frequency, reduced patient anxiety, and improved quality of life ^64^.

Psychological and relational factors also play a role in SSE adherence ^8^. Patients with lower levels of depression, stronger action planning, and well-defined intentions are more likely to consistently engage in SSE. Additionally, partner dynamics is a key moderating factor, as relationship quality can significantly enhance adherence by creating opportunities for one partner to help or remind the other at-risk partner ^170^. Furthermore, self-efficacy scales designed specifically for melanoma patients provide insights into barriers to SSE, emphasizing the influence of physician support, examination intentions, and psychological stress ^160^. Longitudinal studies tracking monthly SSE have identified three distinct patient trajectories: adherent (41%), drop-off (35%), and non-adherent (24%), indicating the need for tailored intervention strategies to support each group ^22^. To improve early detection and long-term outcomes, a multidisciplinary approach integrating educational initiatives, technological solutions, and psychosocial support is essential ^110,181^.

#### Education

Significant gaps in patient education impact melanoma prognosis and patient outcomes. Notably, only 5% of melanoma patients can correctly identify all four key tumor characteristics ^26^. This knowledge gap extends to important medical information: 34% of patients require clarification on their Breslow Tumor Index thickness, 33% do not understand ulceration features, and 65% struggle to understand the specifics of mitosis. These gaps highlight the need for comprehensive, multi-faceted educational programs to improve patient understanding of their condition.

Studies indicate that patients prefer diverse educational formats, with 92% favoring verbal information from physicians, 62% engaging with video resources, and 43% using traditional brochures ^26^. More than three-quarters of patients reported value in YouTube videos on self-inspection of the skin and lymph nodes. These findings suggest that patient education on melanoma and early detection can be significantly improved through accessible digital platforms. There is also a need to enhance educational approaches by complementing information with self-monitoring and early detection methods for possible melanoma changes.

Another important component in improving the prognosis of melanoma patients is enhancing the education of primary care providers (PCPs) ^128,154^. Currently, there is no standardized curriculum for skin cancer examinations and available training programs vary widely in their content, delivery methods, and effectiveness ^154^. However, the INternet curriculum FOR Melanoma Early Detection (INFORMED) program demonstrated a 79% increase in melanoma diagnoses among trained PCPs, showing the promise of educational interventions for healthcare providers ^101^. Beyond medical knowledge, the emotional burden of a melanoma diagnosis and treatment must also be addressed. Nearly 41% of patients report experiencing emotional distress, including worry, fear, and self-consciousness, which can negatively impact their well-being and treatment adherence ^135^. Integrating empathetic, comprehensive education that combines medical information with psychological support can help alleviate this burden.

### Patient/Societal Burden

#### Quality of Life

The current literature on melanoma and melanoma patients has identified numerous gaps in patient quality of life that the healthcare system must address ^74,87^. Some studies have indicated that the development of enhanced predictive prognostic tools could facilitate more informed discussions between healthcare teams and patients/families regarding treatment options and survival outcomes. Additionally, Vogel et al. reported that 50% of study participants felt overwhelmed by the fast pace of melanoma diagnosis and treatment, indicating their psychological care after receiving a potentially life-altering diagnosis was often overlooked ^133^. It should be noted that while the time frame from diagnosis to treatment is rapid, numerous patient-specific factors such as non-white skin, low educational attainment, and non-central residency, have all been found to cause inequitable delays ^13^.

Many melanoma patients report a decreased quality of life due to the overwhelming nature of their diagnosis, surgery/treatment, and recovery process. In particular, there are gaps in psychosocial care, with many patients expressing frustration and anger over being dismissed by healthcare professionals regarding melanoma concerns. This lack of support often leaves patients unprepared for the sudden diagnosis that follows. Rogiers et al. reported that 53% of melanoma survivors experienced clinical levels of anxiety and/or depression ^74,133^. Despite advancements in melanoma treatment, studies show that many patients continue to struggle with post-treatment issues such as anxiety, pain, and subjective cognitive impairment.

#### COVID-19 Impact

The COVID-19 pandemic brought the majority of world sectors to an abrupt halt. As such, many lost their access to non-critical or urgent care. The literature on the impact of the pandemic on melanoma diagnosis and treatment is largely consistent, with studies reporting a sharp and unexpected decline in the rate of melanoma diagnoses and treatments immediately following the implementation of lockdown measures. Specifically, melanoma biopsy and excision rates dropped by 27% and 20%, respectively ^9,33^. This decline was followed by an unprecedented rise in advanced-stage melanoma diagnoses, with lesions found to be thicker than expected. This is significant, as advanced-stage disease and thicker lesions are associated with poorer prognoses and reduced survival rates. Gualdi et al. found that, on average, melanoma lesions increased in thickness by 0.4-0.5 mm during the pandemic, a significant negative prognostic factor ^37^. Experts believe that the COVID-19 pandemic has also contributed to a bottleneck phenomenon, where delays in early screening and access to timely primary and specialized care will result in an ongoing increase in diagnoses of advanced-stage cancers.

### Implications and Limitations

This review examined 183 studies focusing on melanoma patients, healthcare perspectives, and gaps or solutions in patient care, revealing significant deficiencies across multiple areas including intersectionality, treatment, diagnosis/prognosis, and patient/societal burden. The literature demonstrates profound variations in the impact of melanoma on factors such as socioeconomic status, education, age, gender, rural residency, and psychosocial conditions. Thus, our review has wide-reaching implications for the healthcare sector, from policy-making and research funding to clinical practices and patient outreach. The gaps identified in this review suggest urgent areas for future research, particularly in exploring novel, patient-centered approaches to address disparities in melanoma care and psychological support for all melanoma patients. This review could not describe the impact of accessibility, LGBTQ+, cultural, and Indigenous disparities on melanoma outcomes due to a lack of relevant research, which constitute other areas of future research.

Given the broad nature of this review and the lack of quantitative data, individual findings should be interpreted with caution. Furthermore, although numerous factors were considered in this review, the generalizability of these findings is limited, as the review focused on regions with healthcare systems similar to those in Canada and the United States. Additionally, most of the studies meeting inclusion criteria were observational, and as a result, establishing causative effects may be challenging due to the small number of RCTs. Finally, our review incorporated various literature reviews, but it was ensured that those would not be the sole source of information for any observations. However, it is important to note that some degree of bias may still have resulted from the inclusion of these sources.

## CONCLUSION

In this review, gaps in the melanoma patient’s journey from diagnosis onward were analyzed. Through a thorough search of the literature, followed by critical appraisal and analysis of the relevant studies and reviews, four domains of concern were identified: intersectionality, treatment, diagnosis/prognosis, and patient/societal burden. Each domain presents significant challenges for melanoma patients. It is imperative that clinicians, policymakers, and researchers collaborate to address these issues, ensuring the promotion of sustainable, long-term outcomes for all melanoma patients.

## Data Availability

All data produced in the present study are available upon reasonable request to the authors

## STATEMENTS AND DECLARATIONS

## Authorship Contribution

JX devised the project, wrote the protocols, and conducted the search strategy. YCJ conducted the hand-search. JX and AA supervised project completion. JC and AA completed the majority of the title/abstract and full-text screening, with help from YCJ, AD, and YM. AZO and AA extracted and assessed the quality of the majority of included studies, with help from AD, YCJ, JC, YM, and AX. The methods were primarily completed by JC and AX. The introduction, results, implications/limitations, and conclusion were primarily written by AA. Discussions regarding intersectionality were primarily analyzed by JC and AX. Discussions regarding treatment were primarily analyzed by YCJ. Discussions regarding diagnosis/prognosis were primarily analyzed by AD and AZO with help from YCJ. Discussions regarding patient/societal burden were primarily analyzed by YM. All authors contributed to the editing and review process, with a specific focus from YM, AA, and JC.

